# CoViD-19 outbreak in Northern Italy: Did particulate matter really play a key role?

**DOI:** 10.1101/2020.06.11.20128215

**Authors:** Maria Cristina Collivignarelli, Alessandro Abbà, Francesca Maria Caccamo, Giorgio Bertanza, Roberta Pedrazzani, Marco Baldi, Paola Ricciardi, Marco Carnevale Miino

**Author notes:** Corresponding author -> Email address (Marco Carnevale Miino).

## Abstract

Northern Italian regions have been the most affected from CoViD-19 compared to other Italian areas and are also the zones where air pollutants concentration has been higher than in the rest of Italy. The aim of the research is analysing possible correlations between air pollutants PM_10_ and PM_2.5_ and the rapidity of the spread of the infection caused by CoViD-19 in Northern Italy. PM_10_ and PM_2.5_ data for all the 41 studied cities were collected from the local environmental protection agencies. In order to compare air quality data with epidemiological data (T_d_), a statistical analysis was conducted identifying the correlation matrices of Pearson and Spearman, considering the possible incubation period of the disease. The results exclude a strong direct correlation between PM in the air and the diffusion rate of CoViD-19. Further developments are necessary for a better comprehension of the influence of atmospheric pollution parameters on the rapidity of spread of the virus SARS-CoV-2, since a synergistic action with other factors, such as meteorological factors, could not be excluded.

## 1. Introduction

Northern Italy, which has been the most affected area by coronavirus disease (CoViD-19) (INCP, 2020), is also the portion of the Country with the highest amount of atmospheric particulate matter (PM) often exceeding the legislative limit. Moreover, together with Poland and Bulgaria, Northern Italy has the worst air quality in Europe in terms of PM (EEA, 2019, 2018, 2017).

PM is a mixture of solid and liquid particles in the air. Based on its size, PM can be classified into three categories: coarse (2.5–10μm), fine (<2.5μm), and ultrafine (<0.1μm) (Ciencewicki and Jaspers, 2007). Several studies confirmed a strong correlation between air particulate pollution and the increase of respiratory diseases (Cruz-Sanchez et al., 2013; Horne et al., 2018; Xu et al., 2016; Zhou et al., 2015).

The Italian Society of Environmental Medicine (SIMA) (SIMA, 2020) supposed for the first time a possible correlation between the great diffusion of CoViD-19 in Northern Italy and the high concentrations of PM_10_ and PM_2.5_ considering the atmospheric particulate matter as a vector that could transport SARS-CoV-2 (Setti et al., 2020b, 2020a). Several other authors supposed that PM could acts as a support for novel SARS coronavirus (SARS-CoV-2), allowing the spread and the transport even for significant distances. As reported in these studies, PM may represent a substrate that allows the virus to remain in the air in a contagious form for hours or days, promoting its diffusion (Sanità di Toppi et al., 2020; SIMA, 2020). Many studies have been carried out on the association between the concentration of different air pollutants (i.e. PM_2.5_, PM_10_, SO_2_, CO, NO_2_, and O_3_) in the air and the proliferation and aggressiveness of the novel coronavirus infection (Coccia, 2020; Liu et al., 2020; Yari and Moshammer, 2020; Zhu et al., 2020).

This study aims to explore the relationship between ambient air pollutants PM_10_ and PM_2.5_ and the rapidity of the spread of the infection caused by CoViD-19 in Northern Italy, where air pollutants concentration is higher than in the rest of Italy. Data on particulate matter (PM_10_ and PM_2.5_) were analysed and, considering an incubation time of 10 - 15 d, were compared with the CoViD-19 rapidity of spread, evaluated using the doubling time (T_d_), in 41 cities of Northern Italy. In order to compare air quality data with epidemiological data, a statistical analysis was conducted.

## 2. Methods

### 2.1. Area of the study

Considering that CoViD-19 has broken out in Northern Italy, this part of the Country has been selected in order to detect a possible correlation between epidemic spread and PM in air. According to the most recent available data (1.1.2020), the area of the study was larger than 100,000 km^2^ (ISTAT, 2020a) and divided in 41 provinces in seven different Regions (Piedmont, Valle d’Aosta, Lombardy, Liguria, Veneto, Trentino, and Emilia Romagna). The analysis has been applied on the capital of each province, totally accounting for around 25.8 million of inhabitants (ISTAT, 2020b). In Fig. 1, the map of the selected provinces and the location of capital cities are reported.

**Fig 1:**
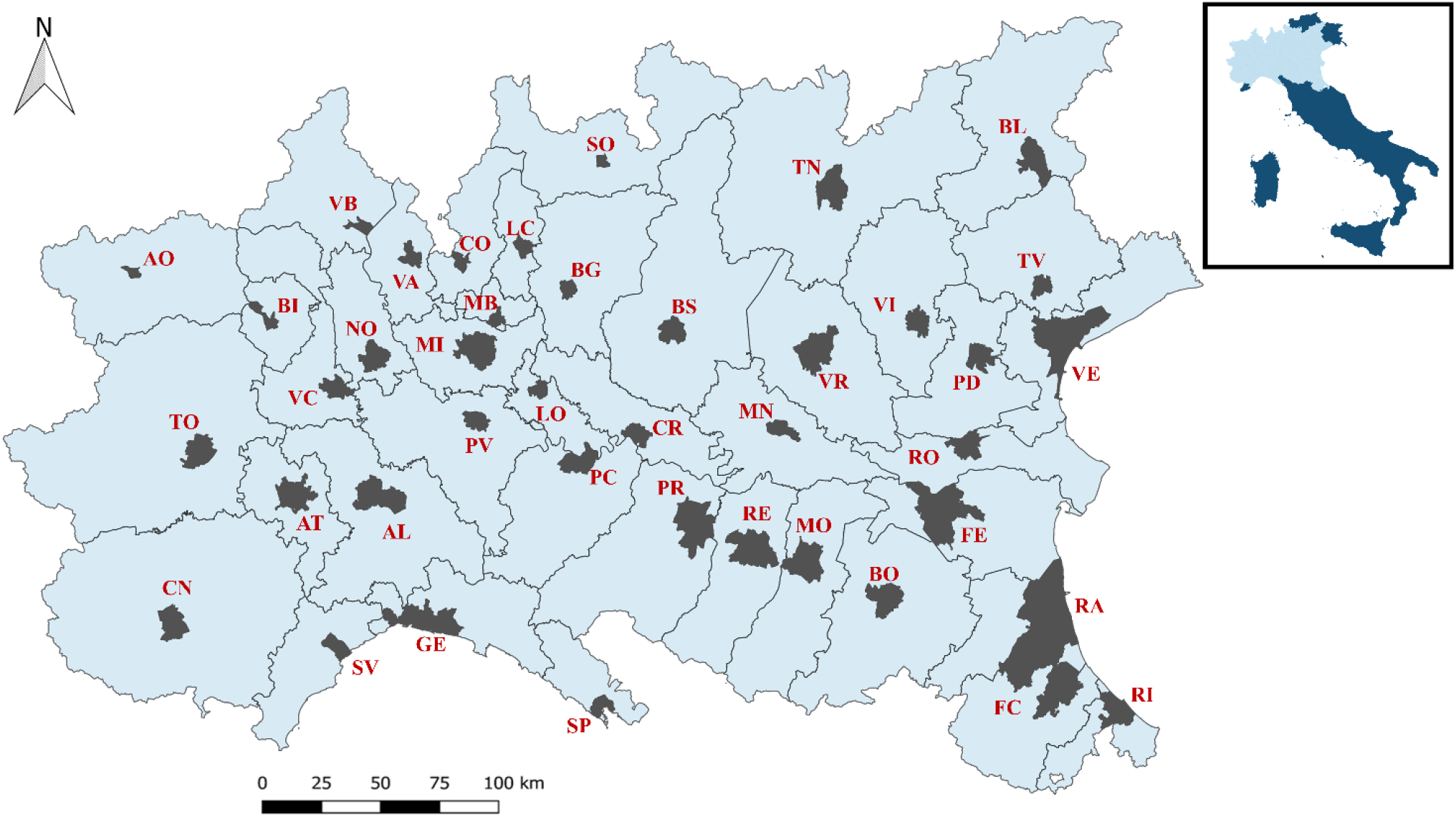
Map of the area analysed in the study and its location in Italy. The capital city for each province is highlighted. Map realized with QGIS (2020). AL: Alessandria; AO: Aosta; AT: Asti; BG: Bergamo; BI: Biella; BL: Belluno; BO: Bologna; BS: Brescia; CN: Cuneo; CO: Como; CR: Cremona; FC: Forlì and Cesena; FE: Ferrara; GE: Genoa; LC: Lecco; LO: Lodi; MB: Monza; MI: Milan; MN: Mantova; MO: Modena; NO: Novara; PC: Piacenza; PD: Padua; PR: Parma; PV: Pavia; RA: Ravenna; RE: Reggio Emilia; RI: Rimini; RO: Rovigo; SO: Sondrio; SP: La Spezia; SV: Savona; TN: Trento; TO: Turin; TV: Treviso; VA: Varese; VB: Verbania; VC: Vercelli; VE: Venice; VI: Vicenza; VR: Verona. [two-column fitting image]

### 2.2. Epidemiological data collection and processing

The epidemiological evolution of the new SARS-CoV-2 cases was not available at city-level. Therefore, the epidemiological data referred to each single province, provided by the Italian National Civil Protection (INCP, 2020), were considered. In order to compare the spread of the CoViD-2019, the new cases in the selected periods of CoViD-19 outbreak were fitted with exponential curve (Eq. 1) aiming to quantify the T_d_ (Eq. 2):

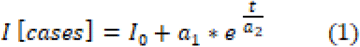

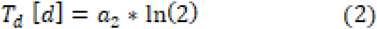

Where *I* represents the number of new infections, and *t* is the time from the first proclaimed case in the province [d].

### 2.3. Particulate matter data collection and processing

The particulate matter, with a diameter of <10 μm (PM_10_) and 2.5 μm (PM_2.5_), data for all cities, were collected from the local environmental protection agencies (APPA Trento, 2020; ARPA Emilia-Romagna, 2020; ARPA Liguria, 2020; ARPA Lombardia, 2020; ARPA Piemonte, 2020; ARPA Valle d’Aosta, 2020; ARPA Veneto, 2020). All air quality control units, located in the capital cities, which measured particulate matters were selected and employed (Table S1) in the selected periods. Forlì and Cesena are co-capitals of their province and were considered as a single city. Data of PM_2.5_ in Belluno, Ferrara and Reggio Emilia were not available. The daily averages (24 h) of the air pollutants for each city were calculated with the median, the standard deviation and the confidence interval.

### 2.4. Comparison of the data

In order to compare air quality data (PM_10_ and PM_2.5_) with epidemiological data (T_d_), a statistical analysis was conducted in order to identify the correlation matrices of Pearson and Spearman. Moreover, three different fittings (linear, quadratic and cubic) were used to investigate a correlation between the CoVid-19 spread rapidity in Northern Italy and particulate matter.

### 2.5. Determination of periods

The choice of the periods determined to select the epidemiological and the air quality data has been made considering the average incubation time of the SARS-CoV-2 in order to evaluate the actual period, during which contagion among people might occur. Several studies determined that the incubation time could be up to 10 - 15 d (Backer et al., 2020; Lai et al., 2020; Li et al., 2020). Therefore, while epidemiological data were considered from the day on which the first person infected in each province was identified, the air quality period was chosen in such a way that it was anticipated by 15 d. In the 8^th^ March, 2020 several restrictions were imposed in part of Northern Italy and in 9^th^ March, 2020 they were extended at the rest of the Country (DPCM, 2020a, 2020b). Following the further increase in the number of infections, the restrictions were made even more severe starting from March 11^th^, 2020 (DPCM, 2020c). In order to study only the exponential tract of the epidemiological curve, no data after the 18^th^ March 2020 were considered (Fig. 2). In some red areas (around the city of Lodi) the restrictions have been imposed earlier than in the rest of the region. The different lockdown timing could have direct repercussions on the epidemiological curve and therefore in Lodi, the selected period ended in 1 March, 2020 (IMH, 2020a, 2020b). In Bergamo and Lecco, the influence due to lockdown were already visible before the 18^th^ March 2020. In these cases, the selected periods were shortened.

**Fig 2:**
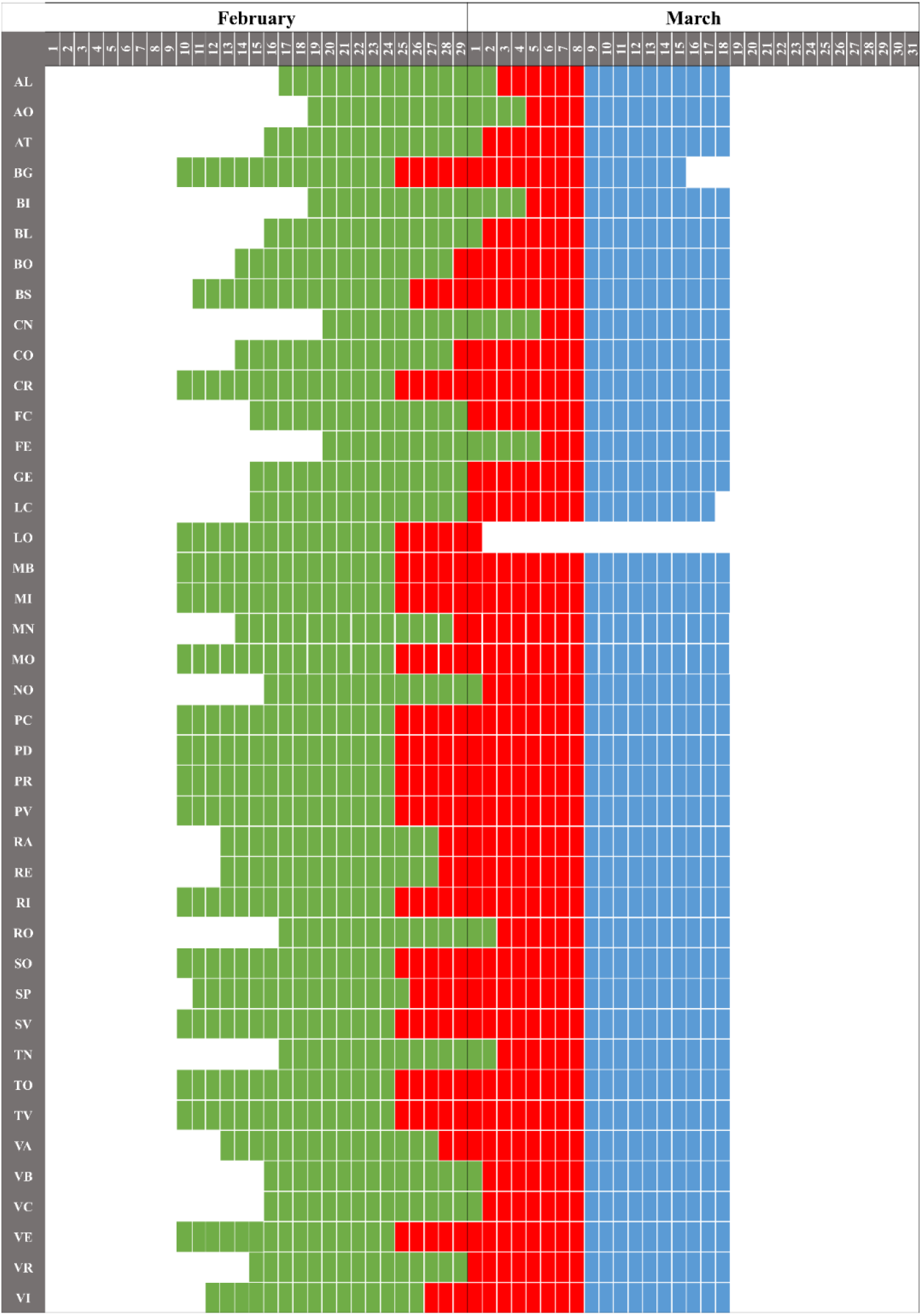
Selected periods for air quality monitoring (green) and epidemiological data (blue) collection. In the periods highlights in red both types of data have been studied. AL: Alessandria; AO: Aosta; AT: Asti; BG: Bergamo; BI: Biella; BL: Belluno; BO: Bologna; BS: Brescia; CN: Cuneo; CO: Como; CR: Cremona; FC: Forlì and Cesena; FE: Ferrara; GE: Genoa; LC: Lecco; LO: Lodi; MB: Monza; MI: Milan; MN: Mantova; MO: Modena; NO: Novara; PC: Piacenza; PD: Padua; PR: Parma; PV: Pavia; RA: Ravenna; RE: Reggio Emilia; RI: Rimini; RO: Rovigo; SO: Sondrio; SP: La Spezia; SV: Savona; TN: Trento; TO: Turin; TV: Treviso; VA: Varese; VB: Verbania; VC: Vercelli; VE: Venice; VI: Vicenza; VR: Verona. (For interpretation of the references to colour in this figure, the reader is referred to the Web version of this article). [two-column fitting image]

The air quality data of PM_10_ and PM_2.5_ were analysed since the 15 day before the date of the first confirmed infection up to the partial lockdown date of 8 March (DPCM, 2020a), as showed in Fig.2. The choice of the period for determine the air quality was made considering the potential incubation time of the novel coronavirus around 14 days (as a maximum precautionary limit) and the restriction imposed on March 8 to evaluate the real period in which there was possible contagion among people.

## 3. Results and discussion

### 3.1. Epidemiological analysis

Fitting the epidemiological data of total infection by an exponential curve (Fig. 3 – Tab. S2), the T_d_ has been calculated for each city considered in the study (Fig.4). This value varied significantly. Asti and Piacenza showed the higher T_d_, 14.9 d and 10.7 d respectively, representing a slower spread of the CoViD-19 infection among the population. On the contrary, other cities such as Lodi, Aosta, Novara and Turin were characterized by a T_d_ fewer than 2.5 d. In these cases, the transmission of the virus among the population was faster. As expected, the minor T_d_ (1.2 d) belongs to Lodi which was the first area of contagion and outbreak of the CoViD-19 in Italy. The T_d_ obtained in this study are in accordance with other results reported in scientific literature. D’Arienzo and Coniglio (2020) identify T_d_ equals to 3.1 d for Italy in the period February 25^th^–March 12^th^, 2020. In the period February 20^th^-March 24^th^, in the Italian regions of Lombardy and Emilia Romagna, Riccardo et al. (2020) evaluated T_d_ equals to 2.7 d. Setti et al. (2020b) highlighted that in Milan, before March 13^th^, T_d_ was 2.0 d.

**Fig 3:**
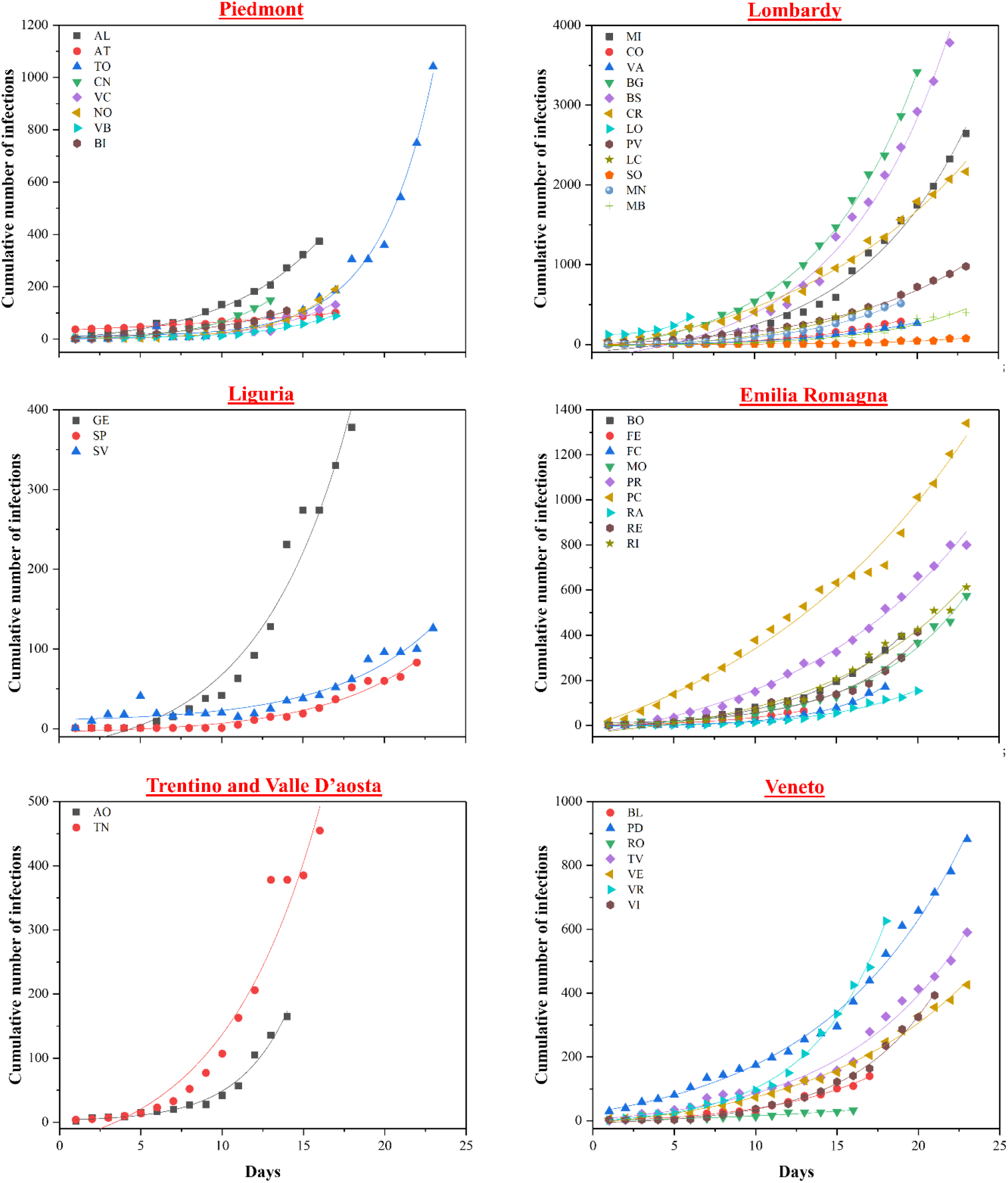
Epidemiological data and exponential fitted curves. AL: Alessandria; AO: Aosta; AT: Asti; BG: Bergamo; BI: Biella; BL: Belluno; BO: Bologna; BS: Brescia; CN: Cuneo; CO: Como; CR: Cremona; FC: Forlì and Cesena; FE: Ferrara; GE: Genoa; LC: Lecco; LO: Lodi; MB: Monza; MI: Milan; MN: Mantova; MO: Modena; NO: Novara; PC: Piacenza; PD: Padua; PR: Parma; PV: Pavia; RA: Ravenna; RE: Reggio Emilia; RI: Rimini; RO: Rovigo; SO: Sondrio; SP: La Spezia; SV: Savona; TN: Trento; TO: Turin; TV: Treviso; VA: Varese; VB: Verbania; VC: Vercelli; VE: Venice; VI: Vicenza; VR: Verona. (For interpretation of the references to colour in this figure, the reader is referred to the Web version of this article). [two-column fitting image]

**Fig 4:**
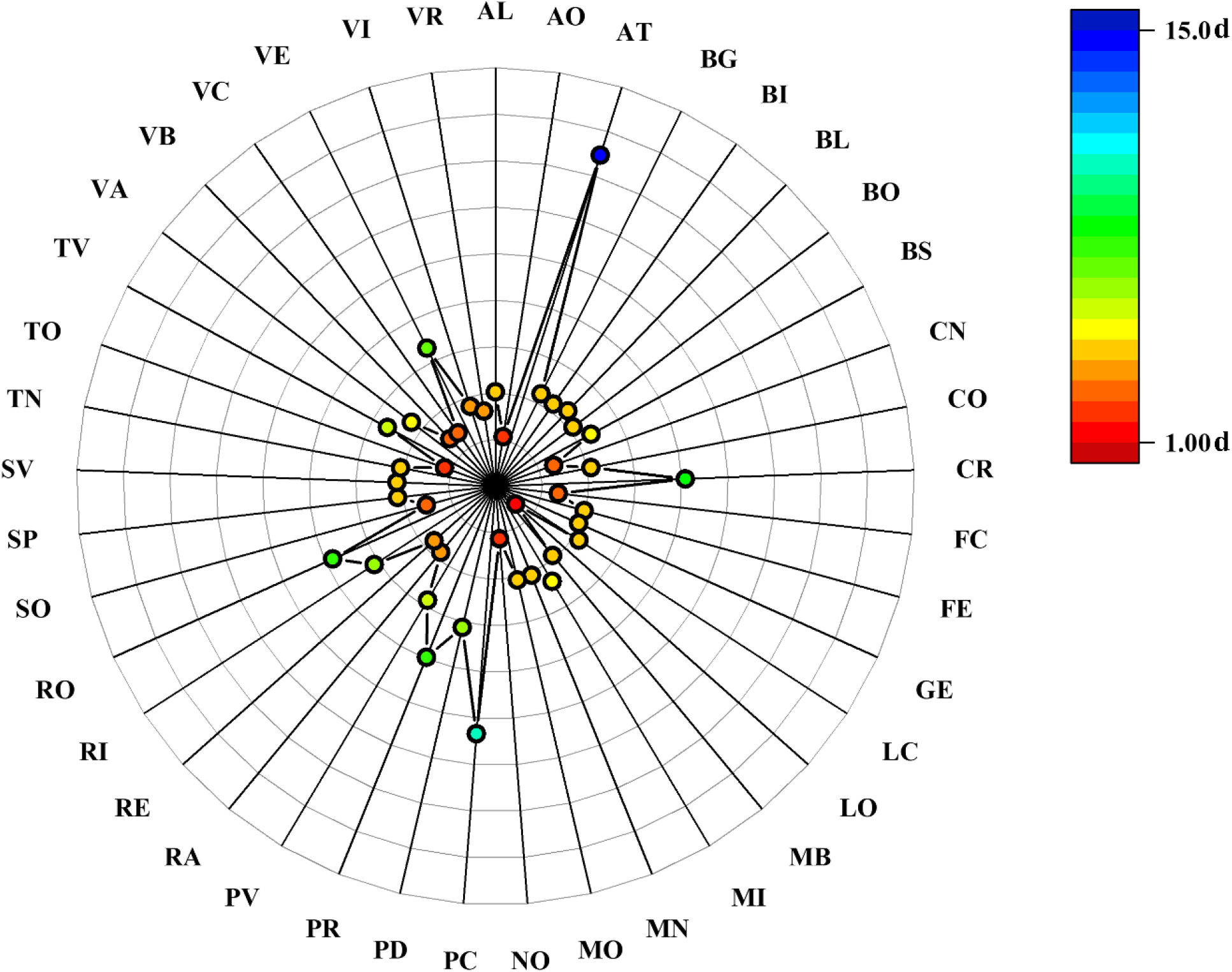
Value of doubling time (T_d_) for each city. AL: Alessandria; AO: Aosta; AT: Asti; BG: Bergamo; BI: Biella; BL: Belluno; BO: Bologna; BS: Brescia; CN: Cuneo; CO: Como; CR: Cremona; FC: Forlì and Cesena; FE: Ferrara; GE: Genoa; LC: Lecco; LO: Lodi; MB: Monza; MI: Milan; MN: Mantova; MO: Modena; NO: Novara; PC: Piacenza; PD: Padua; PR: Parma; PV: Pavia; RA: Ravenna; RE: Reggio Emilia; RI: Rimini; RO: Rovigo; SO: Sondrio; SP: La Spezia; SV: Savona; TN: Trento; TO: Turin; TV: Treviso; VA: Varese; VB: Verbania; VC: Vercelli; VE: Venice; VI: Vicenza; VR: Verona. (For interpretation of the references to colour in this figure, the reader is referred to the Web version of this article). [two-column fitting image]

### 3.2. Particulate matter

In Figure 5, the average and median value of PM_10_ and PM_2.5_ in each city for the selected periods are shown. Among the 41 cities, Cremona, Lodi, Milan, Modena, Padua, Parma, Pavia, Rovigo, Turin, Treviso, Venice and Vicenza presented a mean value of PM_10_ above 40 µg m^-3^. The highest mean value of PM_10_ 48.8 µg m^-3^ was reached in Turin. A similar trend was observed for PM_2.5_, where a concentration higher than 30 µg m^-3^ was found in Cremona, Lodi, Monza, Padua, Pavia, Rovigo, Treviso, Venice and Vicenza. In this case, Padua showed the highest mean value of PM_2.5_ (37.4 µg m^-3^). On the contrary, the lowest mean values of PM_10_ and PM_2.5_, was detected in Aosta, and were equal to 13.8 µg m^-3^ and 9.5 µg m^-3^, respectively. Other areas with the low mean values of PM_10_ and PM_2.5_ were recorder in the seaside cities of Genoa (20.6 µg m^-3^ and 11.8 µg m^-3^, respectively), Savona (20.6 µg m^-3^ and 12.5 µg m^-3^, respectively), and La Spezia (21.2 µg m^-3^ and 10.3 µg m^-3^, respectively), also in this case probably due to weather conditions, such as wind and precipitation, that could have positively influenced the air quality. The concentration of atmospheric PM is highly sensitive to weather conditions and factors such as wind and precipitation can strongly influence its concentration in the air (Baklanov et al., 2016; Collivignarelli et al., 2020).

**Fig 5:**
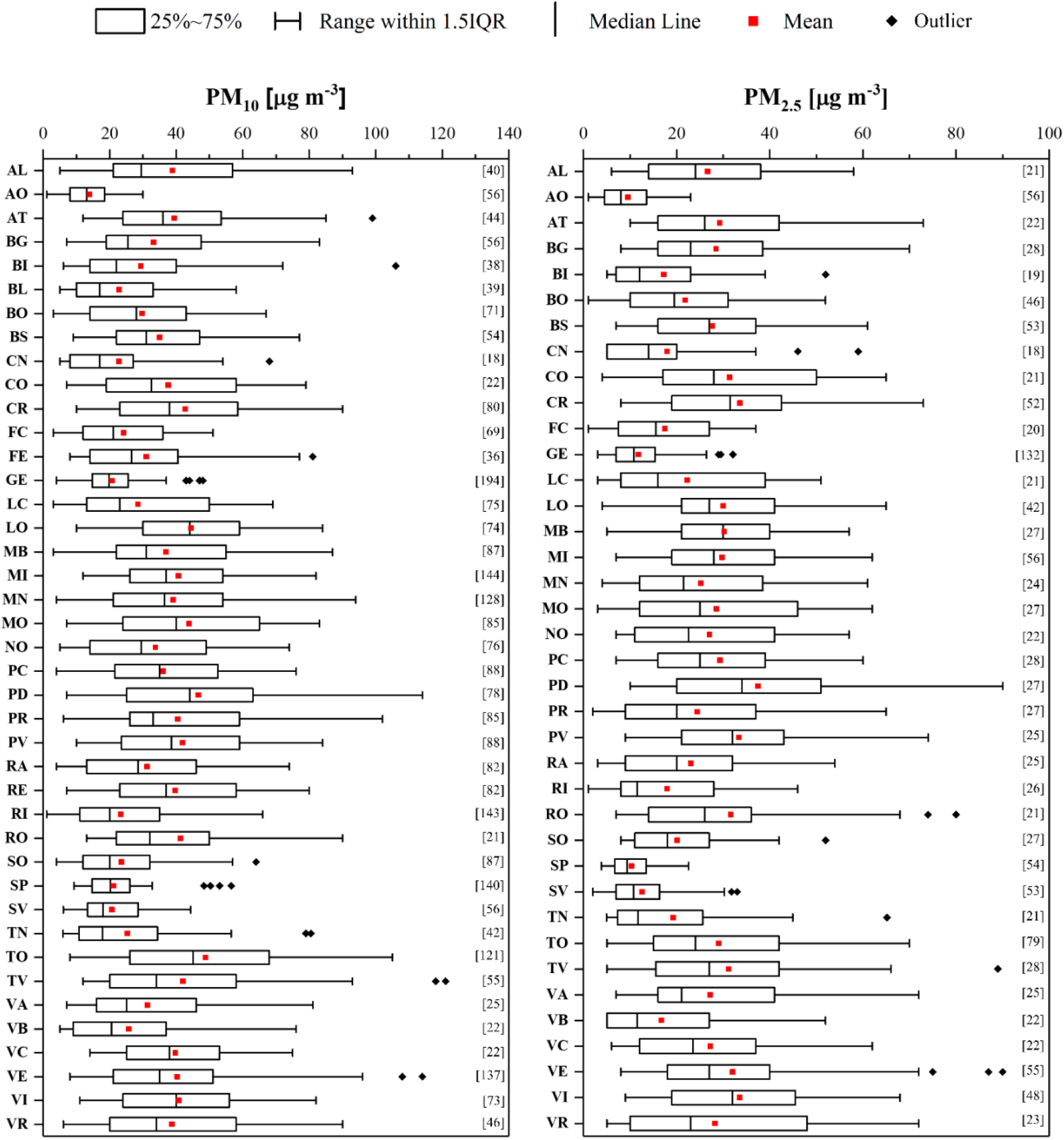
PM_10_ and PM_2.5_ concentration during air quality monitoring period for each city. In brackets, numbers of data are reported. Boxplots represent the distance between the first and third quartiles while whiskers are set as the most extreme (lower and upper) data point not exceeding 1.5 times the quartile range from the median. AL: Alessandria; AO: Aosta; AT: Asti; BG: Bergamo; BI: Biella; BL: Belluno; BO: Bologna; BS: Brescia; CN: Cuneo; CO: Como; CR: Cremona; FC: Forlì and Cesena; FE: Ferrara; GE: Genoa; LC: Lecco; LO: Lodi; MB: Monza; MI: Milan; MN: Mantova; MO: Modena; NO: Novara; PC: Piacenza; PD: Padua; PR: Parma; PV: Pavia; RA: Ravenna; RE: Reggio Emilia; RI: Rimini; RO: Rovigo; SO: Sondrio; SP: La Spezia; SV: Savona; TN: Trento; TO: Turin; TV: Treviso; VA: Varese; VB: Verbania; VC: Vercelli; VE: Venice; VI: Vicenza; VR: Verona. [two-column fitting image]

### 3.3. Statistical analysis and discussion

Air quality and epidemiological data were analysed in order to calculate the Pearson correlation and the Spearman correlation, to assess a possible dependence between the T_d_ of the spread of CoViD-19 among the population and the concentration of PM_10_ and PM_2.5_ in the weeks preceding the identification of the infection (Tab. 1). The results show no significant differences between the two correlations used even if Pearson’s R are slightly higher for T_d_-PM_10_ and PM_2.5_-PM_10_: 0.2485 and 0.8993, respectively. While, as confirmed by the literature (Andrée, 2020), a very strong positive correlation between PM_10_ and PM_2.5_ exists (R = 0.8993 and 0.8669 with Pearson and Spearman, respectively) it is however evident that the correlation between PM and the doubling time of the number of infected people (inversely proportional to propagation speed of the epidemic) is positive and substantially low. In fact, the indices of correlations given by Pearson and Spearman were 0.2485 and 0.1954, respectively.

**Table 1:** Pearson and Spearman correlation for T_d_, PM_10_ and PM_2.5_. a: p-value >0.05. b: p-value < 0.05 [one-column fitting table]

| | | $T_d$ | $PM_{10}$ | $PM_{2.5}$ |
| --- | --- | --- | --- | --- |
| $T_d$ | Pearson | 1 | | |
|  | Spearman | 1 |  |  |
| $PM_{10}$ | Pearson | 0.2485<sup>a</sup> | 1 | |
|  | Spearman | 0.1954<sup>a</sup> | 1 |  |
| $PM_{2.5}$ | Pearson | 0.2995<sup>a</sup> | 0.8993<sup>b</sup> | 1 |
|  | Spearman | 0.2933<sup>b</sup> | 0.8669<sup>b</sup> | 1 |

In order to better study the behaviour of T_d_ as a function of PM_10_ and PM_2.5_, these values have been rented with linear, quadratic and cubic functions (Fig. 6). In the T_d_ -PM_10_ case, the function that best approximates the points is that of 3rd-degree (R^2^ = 0.117), while in the case of T_d_ - PM_2.5_ both the 2nd-degree function and the 3rd-degree function return the best R^2^ (0.109). Two aspects can be highlighted: (i) the 2nd- and 3rd-degree functions do not substantially improve the fitting of the points with respect to the linear function; (ii) in all cases the R^2^ values remain decidedly low in order to define the accurate fitting.

**Fig 6:**
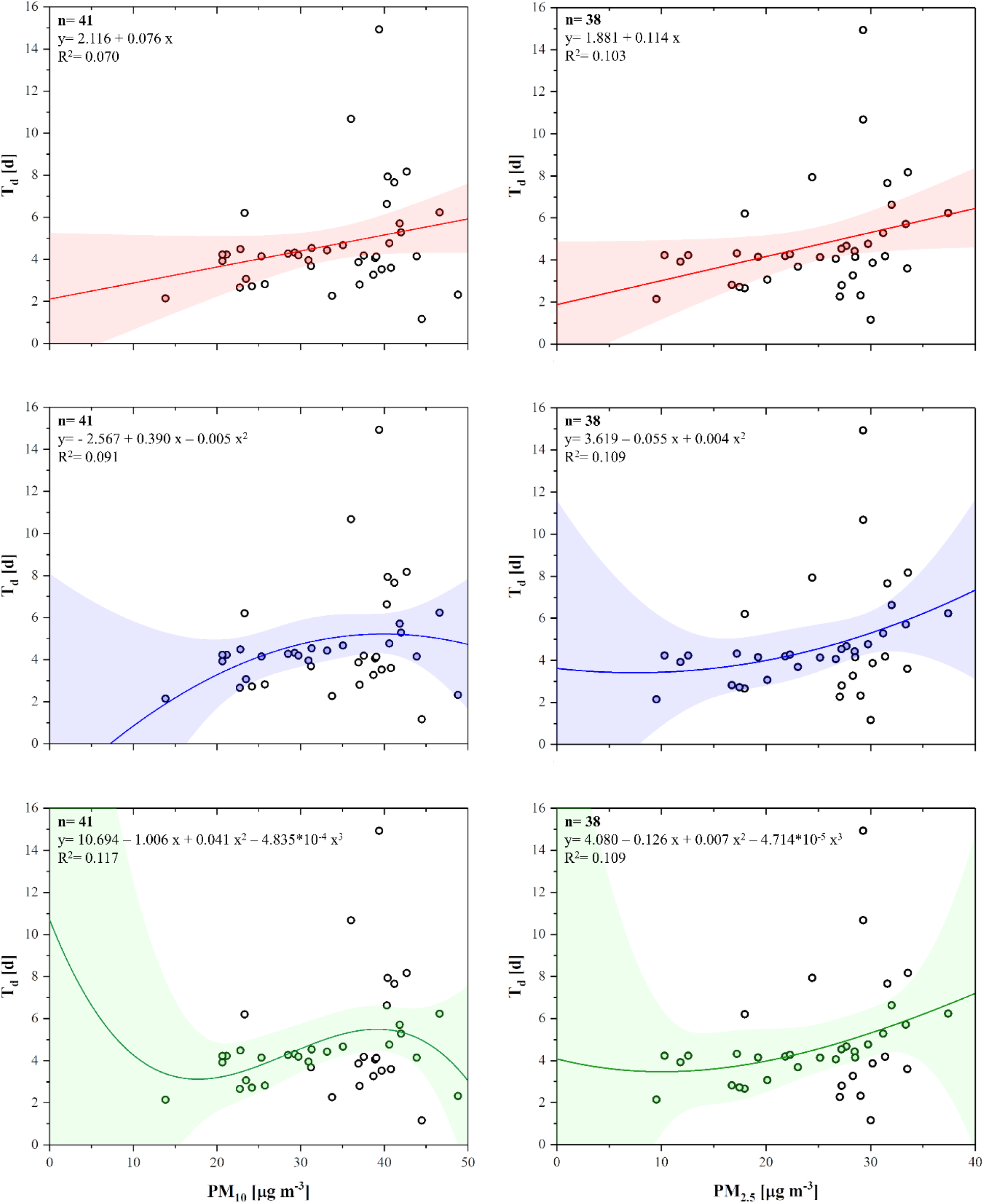
Fitting of T_d_ with PM_10_ and PM_2.5_ with linear (red), parabolic (blue) and cubic (green) functions. The coloured bands represent the 95% confidence interval. n: number of data. (For interpretation of the references to colour in this figure, the reader is referred to the Web version of this article). [double-column fitting table]

Although this study does not consider some factors, such as the possibility that a person may have become infected in a location other than that of residence, it compares the epidemiological and air quality data of 41 cities in Northern Italy in the most affected regions from CoViD-19, taking into account the possible incubation period of the disease. The results allow excluding a strong correlation between the diffusion rate of CoViD-19 and PM. Furthermore, it is interesting to note that the low correlation indices identified are both positive.

This result is only partially comparable with literature results, where in most cases the comparison of PM to the rate of infected people was assessed, instead of the rapidity of contagion, with the use of T_d_. However, the opinion of the scientific community on the influence of PM_10_ on the infected population rate is conflicting. Other studies have instead obtained positive correlations between the number of infected, mortality and the concentration of PM in the air (Ma et al., 2020; Zhu et al., 2020). Anyway, the atmospheric pollution is univocally recognized as a crucial co-factor in worsening the initial health state, inducing the need of intensive care and hospitalization and causing lethality, due to impairment and weakening of the immune system, particularly in case of chronic exposures. Fine and ultrafine particulate matter, as well as ozone, nitrogen dioxide and sulphur dioxide produce a systemic inflammation (with an intense synthesis of proinflammatory cytokines) also in young and non-smoker people, thus deeply increasing the health risks; hypertension, cardiovascular diseases, chronic obstructive pulmonary diseases, diabetes can be listed among the effects caused by air pollution (Conticini et al., 2020; Fattorini and Regoli, 2020; Ogen, 2020).

On the other hand, it is becoming increasingly clear, also by following the epidemic trends worldwide, that several aspects should also be taken into account: focusing exclusively on air pollution can lead to spurious associations, because socio-economic and cultural factors often play a concurrent role. Andree (2020) presents a detailed analysis of the Dutch situation, by studying areas, hotspots, pollution loads, social links, habits, age, gender, household composition, and lifestyle. In addition, the variability in healthcare systems and identification practices highly affect the data about pandemic behaviour.

## 4. Conclusion

In this work, data on the concentration of PM_10_ PM_2.5_ were analysed and, considering an incubation time of 10 - 15 d, were compared with the rapidity of spread of CoViD-19 in 41 cities of Northern Italy. Although this work allows to exclude a strong direct correlation between PM in the air and the diffusion rapidity of CoViD-19, the authors believe it is necessary to further comprehend the influence of atmospheric pollution parameters on the rapidity of spread of the virus SARS-CoV-2, since it cannot be excluded that a synergistic action with other factors, such as meteorological factors, could exist.

## CRediT authorship contribution statement

**Maria Cristina Collivignarelli:** Conceptualization, Methodology, Supervision, Validation, Writing. **Alessandro Abbà:** Methodology, Validation, Visualization, Writing. **Francesca Maria Caccamo:** Data curation and formal analysis, Writing. **Giorgio Bertanza:** Methodology, Validation, Writing. **Roberta Pedrazzani:** Data curation and formal analysis, Validation, Writing. **Marco Baldi:** Visualization, Validation **Paola Ricciardi:** Visualization, Writing. **Marco Carnevale Miino:** Conceptualization, Data curation and formal analysis, Methodology, Supervision, Validation, Writing.

## Data Availability

Nothing to declare.

---

**Table S1:**
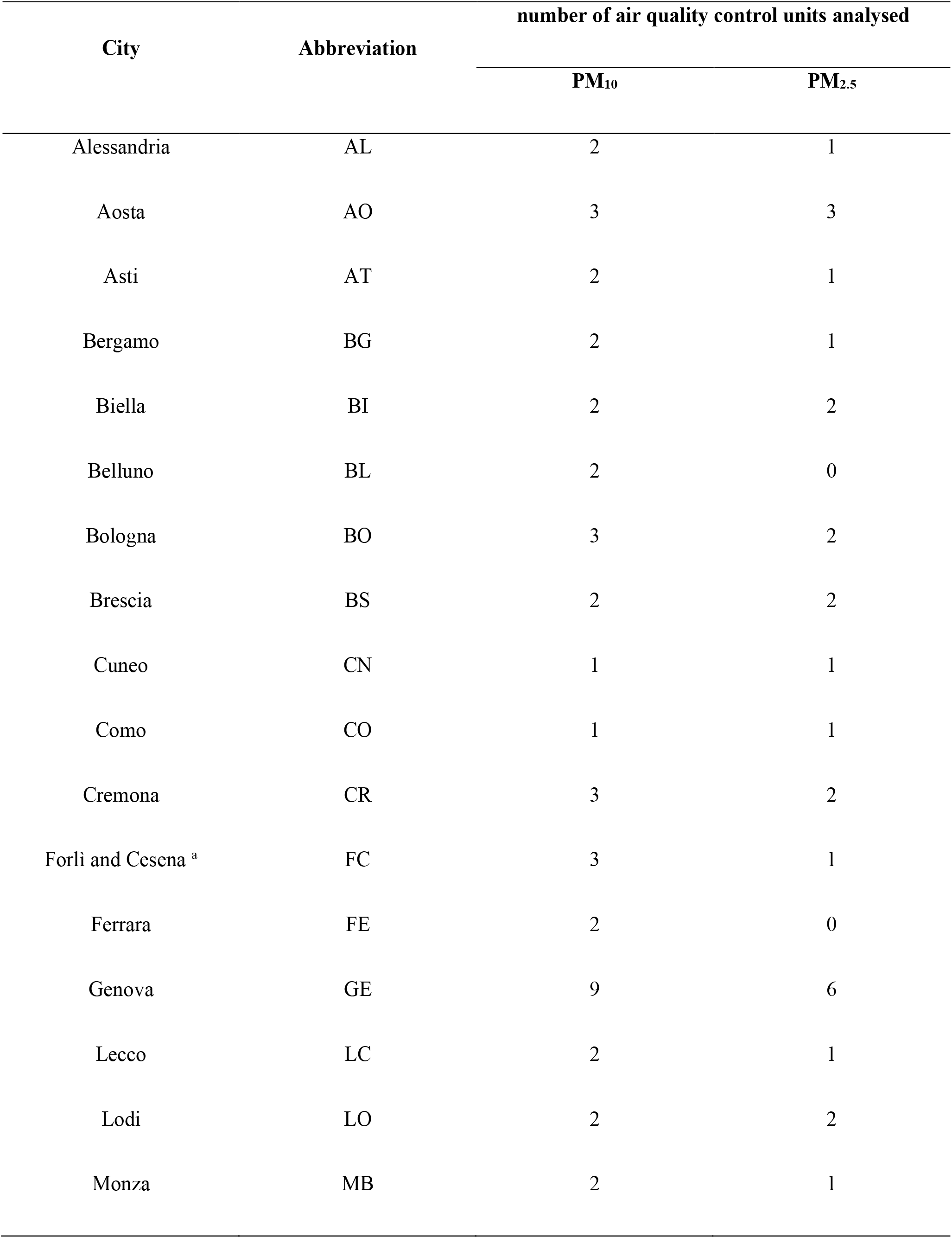
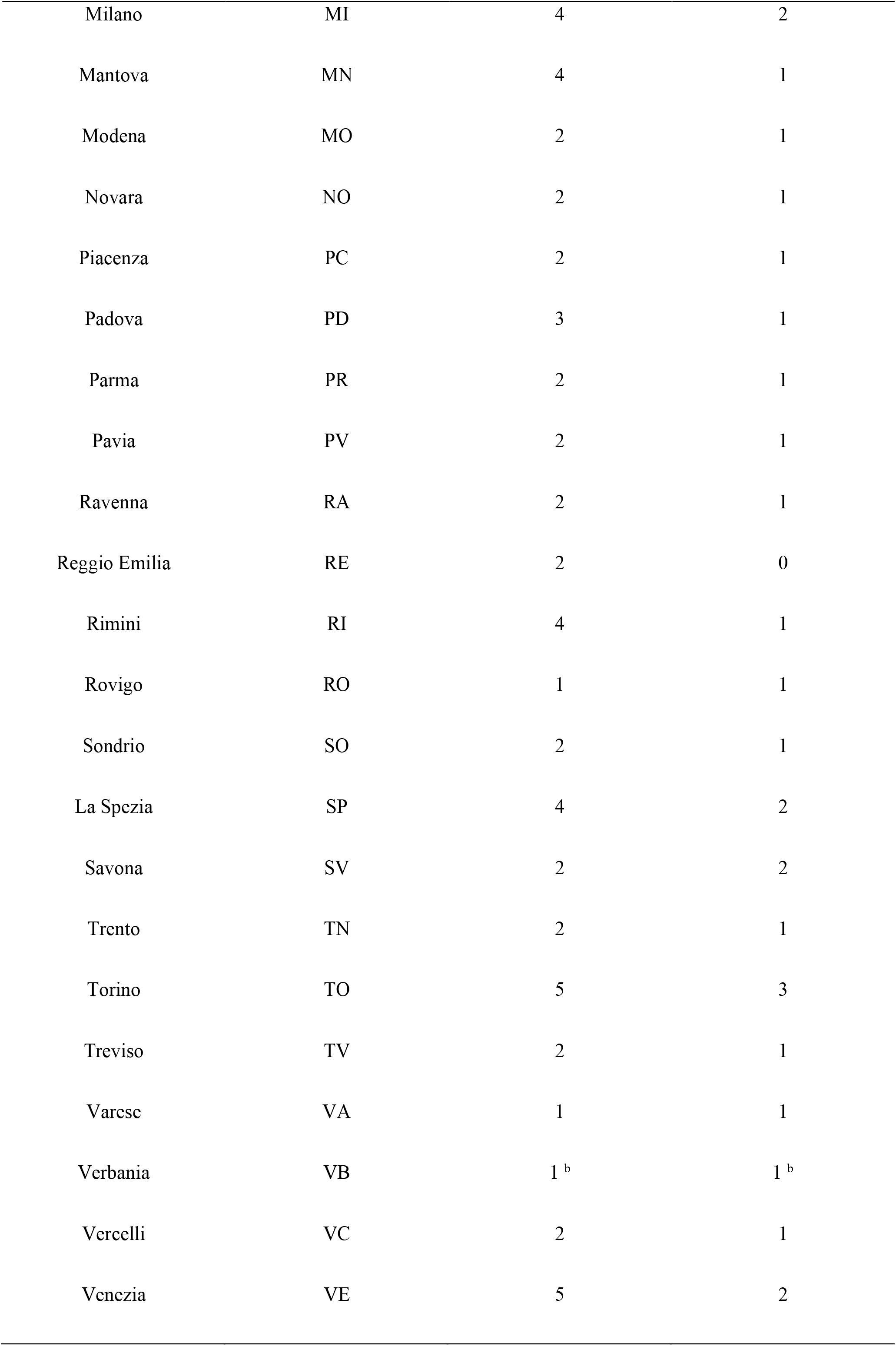
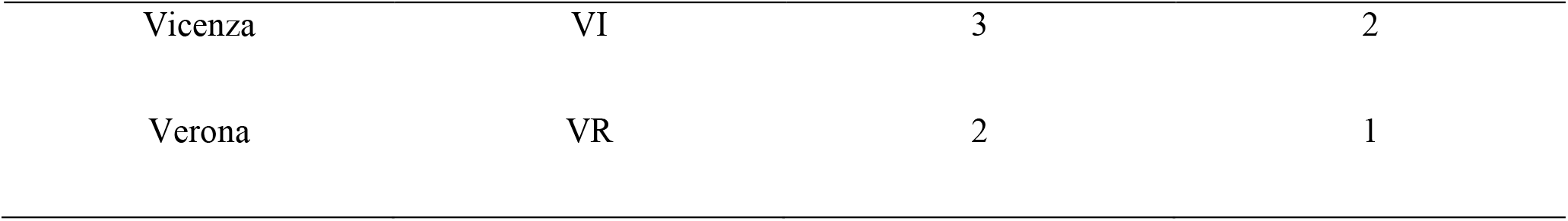
Number of air quality control units for PM_10_ and PM_2.5_ analysed in the study. a: Forlì and Cesena, co-capitals of their province, have been considered as a single city; b: The data refer to the air quality control unit located in Domodossola.

| City | Abbreviation | number of air quality control units analysed |  |
| --- | --- | --- | --- |
| | | $PM_{10}$ | $PM_{2.5}$ |
| Alessandria | AL | 2 | 1 |
| Aosta | AO | 3 | 3 |
| Asti | AT | 2 | 1 |
| Bergamo | BG | 2 | 1 |
| Biella | BI | 2 | 2 |
| Belluno | BL | 2 | 0 |
| Bologna | BO | 3 | 2 |
| Brescia | BS | 2 | 2 |
| Cuneo | CN | 1 | 1 |
| Como | CO | 1 | 1 |
| Cremona | CR | 3 | 2 |
| Forlì and Cesena <sup>a</sup> | FC | 3 | 1 |
| Ferrara | FE | 2 | 0 |
| Genova | GE | 9 | 6 |
| Lecco | LC | 2 | 1 |
| Lodi | LO | 2 | 2 |
| Monza | MB | 2 | 1 |
| Milano | MI | 4 | 2 |
| Mantova | MN | 4 | 1 |
| Modena | MO | 2 | 1 |
| Novara | NO | 2 | 1 |
| Piacenza | PC | 2 | 1 |
| Padova | PD | 3 | 1 |
| Parma | PR | 2 | 1 |
| Pavia | PV | 2 | 1 |
| Ravenna | RA | 2 | 1 |
| Reggio Emilia | RE | 2 | 0 |
| Rimini | RI | 4 | 1 |
| Rovigo | RO | 1 | 1 |
| Sondrio | SO | 2 | 1 |
| La Spezia | SP | 4 | 2 |
| Savona | SV | 2 | 2 |
| Trento | TN | 2 | 1 |
| Torino | TO | 5 | 3 |
| Treviso | TV | 2 | 1 |
| Varese | VA | 1 | 1 |
| Verbania | VB | 1<sup>b</sup> | 1<sup>b</sup> |
| Vercelli | VC | 2 | 1 |
| Venezia | VE | 5 | 2 |
| Vicenza | VI | 3 | 2 |
| Verona | VR | 2 | 1 |

**Table S2:** Values of parameter I_0_, a_1_ and a_2_ of the fitted curve for each city. n: number of epidemiological data used; AL: Alessandria; AO: Aosta; AT: Asti; BG: Bergamo; BI: Biella; BL: Belluno; BO: Bologna; BS: Brescia; CN: Cuneo; CO: Como; CR: Cremona; FC: Forlì and Cesena; FE: Ferrara; GE: Genoa; LC: Lecco; LO: Lodi; MB: Monza; MI: Milan; MN: Mantova; MO: Modena; NO: Novara; PC: Piacenza; PD: Padua; PR: Parma; PV: Pavia; RA: Ravenna; RE: Reggio Emilia; RI: Rimini; RO: Rovigo; SO: Sondrio; SP: La Spezia; SV: Savona; TN: Trento; TO: Turin; TV: Treviso; VA: Varese; VB: Verbania; VC: Vercelli; VE: Venice; VI: Vicenza; VR: Verona.

| | $I_0$ | | $a_1$ | | $a_2$ | | Statistics | | | $n$ |
| --- | --- | --- | --- | --- | --- | --- | --- | --- | --- | --- |
| | Value | Standard Error | Value | Standard Error | Value | Standard Error | Reduced Chi-Sqr | $R^2$ | Adj. $R^2$ | Value |
| AL | -22.7858 | 9.56625 | 26.19837 | 5.03171 | 5.85865 | 0.38588 | 75.97882 | 0.99504 | 0.99427 | 16 |
| AO | 1.71263 | 4.29319 | 1.8668 | 0.83855 | 3.09808 | 0.30763 | 53.16056 | 0.98344 | 0.98043 | 14 |
| AT | -21.7287 | 35.72876 | 55.13212 | 33.69611 | 21.54214 | 9.00583 | 11.14741 | 0.97627 | 0.97288 | 17 |
| BG | -203.838 | 35.39495 | 158.0004 | 15.60995 | 6.38705 | 0.19475 | 2007.19414 | 0.99831 | 0.99811 | 20 |
| BI | -10.3079 | 10.53521 | 12.37323 | 6.7235 | 6.23368 | 1.34012 | 41.51329 | 0.96776 | 0.9619 | 14 |
| BL | -12.0827 | 5.40711 | 10.87774 | 2.98679 | 6.47641 | 0.62986 | 22.7202 | 0.98903 | 0.98746 | 17 |
| BO | -24.3799 | 5.02768 | 18.29187 | 2.20301 | 6.05196 | 0.22422 | 38.719 | 0.99764 | 0.99735 | 19 |
| BS | -327.728 | 87.59519 | 162.9651 | 36.36456 | 6.74469 | 0.44975 | 15296.66559 | 0.99051 | 0.98951 | 22 |
| CN | -6.67553 | 2.70186 | 5.35517 | 1.02009 | 3.83916 | 0.20899 | 9.38547 | 0.99659 | 0.99591 | 13 |
| CO | -25.695 | 7.22943 | 14.06432 | 3.15443 | 6.03549 | 0.41548 | 80.71223 | 0.99188 | 0.99087 | 19 |
| CR | -444.391 | 106.4128 | 388.9752 | 76.50944 | 11.78469 | 1.02075 | 4300.60973 | 0.99211 | 0.99132 | 23 |
| FC | -2.94276 | 1.60517 | 1.82318 | 0.30701 | 3.92652 | 0.14511 | 9.96144 | 0.99673 | 0.99629 | 18 |
| FE | -8.64401 | 3.00631 | 7.68614 | 1.88989 | 5.70994 | 0.5489 | 3.27702 | 0.99388 | 0.99265 | 13 |
| GE | -40.0979 | 20.25716 | 18.40972 | 8.66347 | 5.6552 | 0.80836 | 619.3506 | 0.96761 | 0.9633 | 18 |
| LC | -58.857 | 21.12615 | 33.29357 | 11.01259 | 6.16447 | 0.69497 | 407.23546 | 0.98425 | 0.982 | 17 |
| LO | 112.6962 | 7.52882 | 6.44749 | 2.38324 | 1.67774 | 0.1642 | 34.68078 | 0.99701 | 0.99502 | 6 |
| MB | -18.0619 | 12.94899 | 7.54365 | 3.34796 | 5.57771 | 0.6023 | 654.85582 | 0.96679 | 0.96347 | 23 |
| MI | -193.16 | 48.51016 | 103.1093 | 19.34726 | 6.87684 | 0.37781 | 5278.97352 | 0.99298 | 0.99228 | 23 |
| MN | -35.5755 | 10.91358 | 23.69597 | 4.67477 | 5.96483 | 0.35764 | 190.50716 | 0.99374 | 0.99296 | 19 |
| MO | -11.8609 | 7.01142 | 12.65234 | 2.13228 | 5.98176 | 0.26129 | 161.3006 | 0.99479 | 0.99427 | 23 |
| **NO** | 1.05772 | 2.13991 | 1.07213 | 0.27251 | 3.27579 | 0.16221 | 21.36825 | 0.99404 | 0.99318 | 17 |
| **PC** | -373.065 | 113.1096 | 372.6174 | 92.91388 | 15.40336 | 1.98866 | 1803.72403 | 0.98904 | 0.98794 | 23 |
| **PD** | -46.9666 | 21.64301 | 73.40548 | 12.44327 | 8.99543 | 0.55457 | 455.29133 | 0.99386 | 0.99324 | 23 |
| **PR** | -173.927 | 34.89795 | 138.7576 | 24.62346 | 11.44977 | 0.87783 | 513.0932 | 0.9935 | 0.99285 | 23 |
| **PV** | -61.2791 | 16.11574 | 65.57775 | 8.37606 | 8.23669 | 0.35734 | 336.50171 | 0.99654 | 0.99619 | 23 |
| **RA** | -8.43847 | 3.45396 | 3.89253 | 1.09444 | 5.32099 | 0.39459 | 31.98398 | 0.98769 | 0.98624 | 20 |
| **RE** | 1.80809 | 10.56138 | 7.45922 | 3.03678 | 5.09053 | 0.52539 | 335.14555 | 0.97534 | 0.97243 | 20 |
| **RI** | -85.7284 | 23.86018 | 54.49339 | 13.64294 | 8.94897 | 0.81163 | 562.94886 | 0.9867 | 0.98537 | 23 |
| **RO** | -10.468 | 8.99155 | 10.48627 | 7.42367 | 11.0481 | 4.09031 | 6.98982 | 0.94737 | 0.93927 | 16 |
| **SO** | 0.1461 | 1.82642 | 0.45368 | 0.23829 | 4.43206 | 0.45637 | 21.23177 | 0.96364 | 0.96001 | 23 |
| **SP** | -5.86152 | 2.89049 | 2.48002 | 1.00254 | 6.10327 | 0.67652 | 22.08383 | 0.97105 | 0.96801 | 22 |
| **SV** | 8.74676 | 5.25446 | 2.76366 | 1.66851 | 6.10318 | 0.9723 | 85.98601 | 0.93618 | 0.9298 | 23 |
| **TN** | -68.5405 | 44.61826 | 38.7279 | 24.07553 | 5.98613 | 1.29781 | 1541.30491 | 0.9502 | 0.94254 | 16 |
| **TO** | 10.89431 | 8.37738 | 1.05146 | 0.32278 | 3.35126 | 0.15323 | 681.96806 | 0.9915 | 0.99065 | 23 |
| **TV** | -26.6831 | 15.83901 | 30.54156 | 7.42625 | 7.62375 | 0.59162 | 414.17647 | 0.98779 | 0.98656 | 23 |
| **VA** | -22.8631 | 7.72903 | 14.17442 | 3.53634 | 6.5454 | 0.51432 | 88.87846 | 0.9892 | 0.98793 | 20 |
| **VB** | 1.62903 | 1.69677 | 1.39914 | 0.41722 | 4.07234 | 0.2903 | 8.48801 | 0.98965 | 0.98817 | 17 |
| **VC** | -1.67723 | 5.38567 | 2.11354 | 1.29882 | 4.0414 | 0.58956 | 87.07359 | 0.95775 | 0.95171 | 17 |
| **VE** | -50.8635 | 10.09588 | 44.12839 | 6.16946 | 9.56299 | 0.50902 | 80.64094 | 0.99582 | 0.9954 | 23 |
| **VI** | -13.4788 | 4.33654 | 7.28041 | 1.1698 | 5.19408 | 0.20637 | 63.79283 | 0.99592 | 0.99547 | 21 |
| **VR** | -14.7726 | 7.51136 | 14.06908 | 2.28671 | 4.71441 | 0.19884 | 141.37564 | 0.99641 | 0.99593 | 18 |

